# Predictive mathematical models for the number of individuals infected with COVID-19

**DOI:** 10.1101/2020.05.02.20088591

**Authors:** A.S. Fokas, N. Dikaios, G.A. Kastis

## Abstract

We model the time-evolution of the number *N(t)* of individuals *reported* to be infected in a given country with a specific virus, in terms of a Riccati equation. Although this equation is nonlinear and it contains time-dependent coefficients, it can be solved in closed form, yielding an expression for *N(t)* that depends on a function *α(t)*. For the particular case that *α(t)* is constant, this expression reduces to the well-known *logistic formula*, giving rise to a sigmoidal curve suitable for modelling usual epidemics. However, for the case of the COVID-19 pandemic, the long series of available data shows that the use of this simple formula for predictions underestimates *N(t);* thus, the logistic formula only provides a *lower bound* of *N(t)*. After experimenting with more than 50 different forms of *α(t)*, we introduce two novel models that will be referred to as “*rational”* and *“birational”*. The parameters specifying these models (as well as those of the logistic model), are determined from the available data using an error-minimizing algorithm. The analysis of the applicability of the above models to the cases of China and South Korea suggest that they yield more accurate predictions, and importantly that they may provide an *upper bound* of the actual *N(t)*. Results are presented for Italy, Spain, and France.

## 1. Introduction

The novel coronavirus 2019-nCoV initially emerged in Wuhan, China, at the end of 2019. It is the third coronavirus to appear in the human population in the past two decades, following the severe acute respiratory syndrome coronavirus SARS-CoV outbreak in 2002, and the Middle East syndrome coronavirus MERS-CoV outbreak in 2012. China responded quickly to this outbreak by informing the World Health Organization. Also, after Chinese scientists identified the sequence of the causative virus [1], this information was immediately shared with the international community. Furthermore, China took effective measures for the containment of the spread of this outbreak. The 2019-nCoV is less pathogenic than the earlier two coronaviruses [2]. For example, in the first case of pneumonia caused by this virus reported in USA, a 35-years-old, healthy, individual (who had travelled in Wuhan) presented in a hospital four days after he experienced dry cough and low grade fever; he proceeded to develop pneumonia 5 days later, but quickly recovered [3]. This is the typical disease course for young persons; however, the 2019-nCoV has a significant mortality rate for elderly persons and for those with a variety of underlying medical conditions, including respiratory and cardiovascular diseases, as well as diabetes mellitus. Furthermore, this virus is highly contagious. As a result of these facts, and the lack of appropriate *early* international measures for the suppression of its spread, it has now caused a pandemic.

This pandemic represents the most serious global public health threat since the devastating 1918 H1N1 influenza pandemic. Justifiably, several countries have adopted draconian measures to combat this threat. The scientific community, in addition to its accelerated efforts to develop an effective treatment and a vaccination, is also playing an important role in advising policy makers of possible non-pharmacological approaches to limit the catastrophic impact of the pandemic. For example, two possible strategies, called mitigation and suppression, are thoroughly discussed in the important paper [4]; in the early stages of the pandemic, UK was following mitigation, but after the publication of this report, is now following suppression.

The main goal of this paper is to develop suitable mathematical models for *predicting the time evolution of the cumulative number, N(t), of the individuals reported to be infected, in a given country, by COVID-19*. These models can be used for predicting several features of the epidemic, such as the time that a plateau will be reached, as well as the total number of individuals reported to be infected at that time. Here, *the plateau is defined as the time when the rate of change of the people reported to be infected is 5% of the maximum rate of infection*. We introduce two novel models, called “*rational”* and “*birational*”; the important advantage of these models is that they provide more accurate predictions for the characteristics of the plateau. Also, importantly, the birational model may provide an upper bound of *N(t)*, and hence it is preferable to the rational one. However, the rational model can be constructed sooner than the birational one: the input needed for the rational model is data until around the time, T, when the maximum rate of infection occurs (the corresponding point on the curve describing *N(t)* is known as the *inflection point)*. On the other hand, the birational model requires data for several more additional days. *Assuming that the number of individuals reported to be infected is a time-invariant percentage of the actual number of infected persons*, the model discussed here should be useful for long-term planning strategy and for reassuring the public.

A prerequisite for the development of any accurate model is the existence of appropriate data. For the pandemic of COVID-19, such data are already available: There exist a long series of data from China and South Korea, where their COVID-19 epidemics are passed the plateau. Italy, Spain, and France passed the inflection point several days ago, and UK has now also passed this point. The total reported cases of the above countries (as well as of Greece where the situation is better and USA where it is worse) are shown in figure 1. Estimated rates of change (new reported cases) for Italy, Spain, France, and UK are plotted in figure 2.

**Figure 1.**
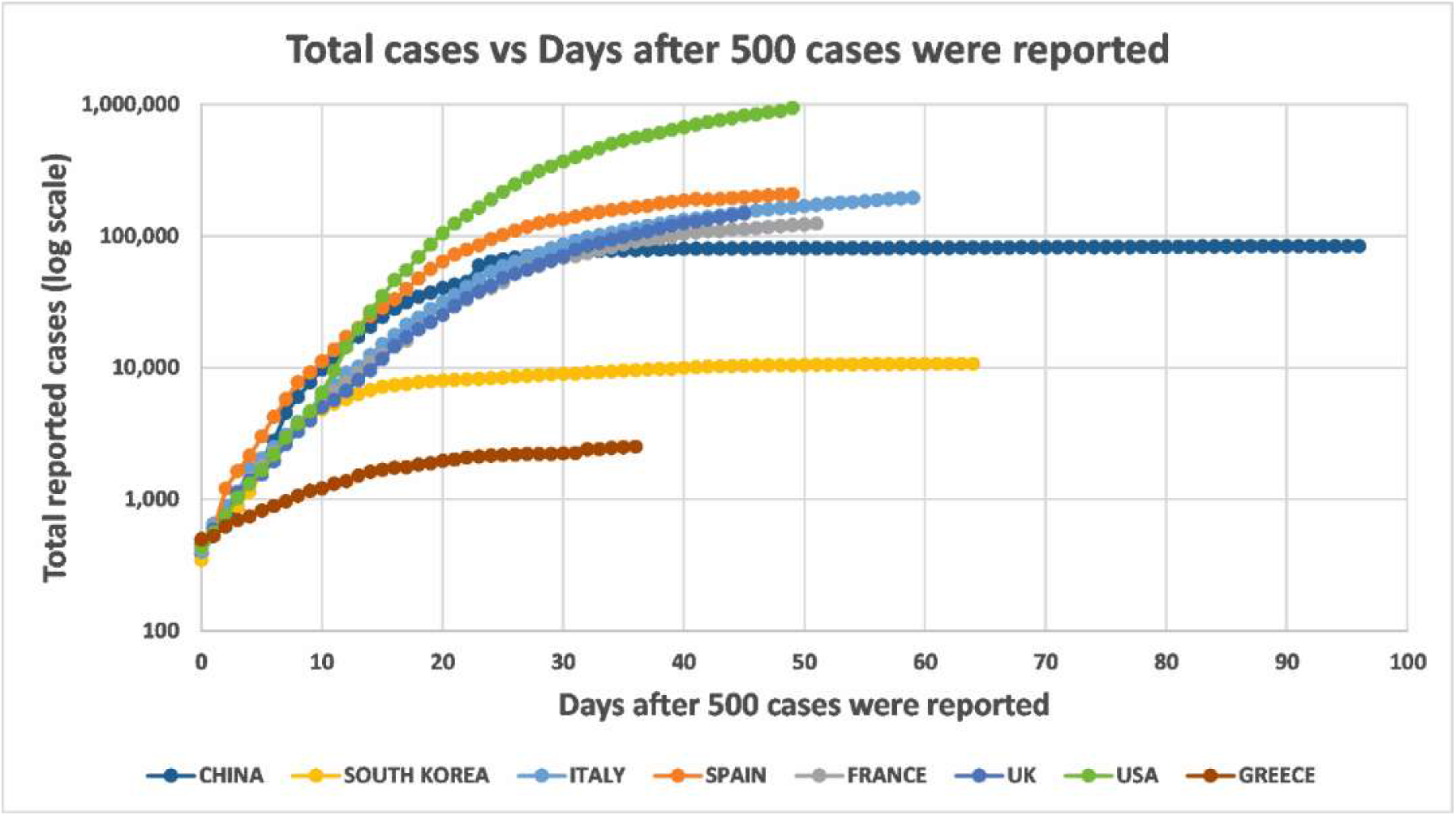
COVID-19 virus epidemics in China, South Korea, Italy, Spain, France, UK, USA, and Greece: Total cumulative number of individuals reported to be infected up to April 26, 2020, as a function of days after the day that 500 cases were reported.

**Figure 2.**
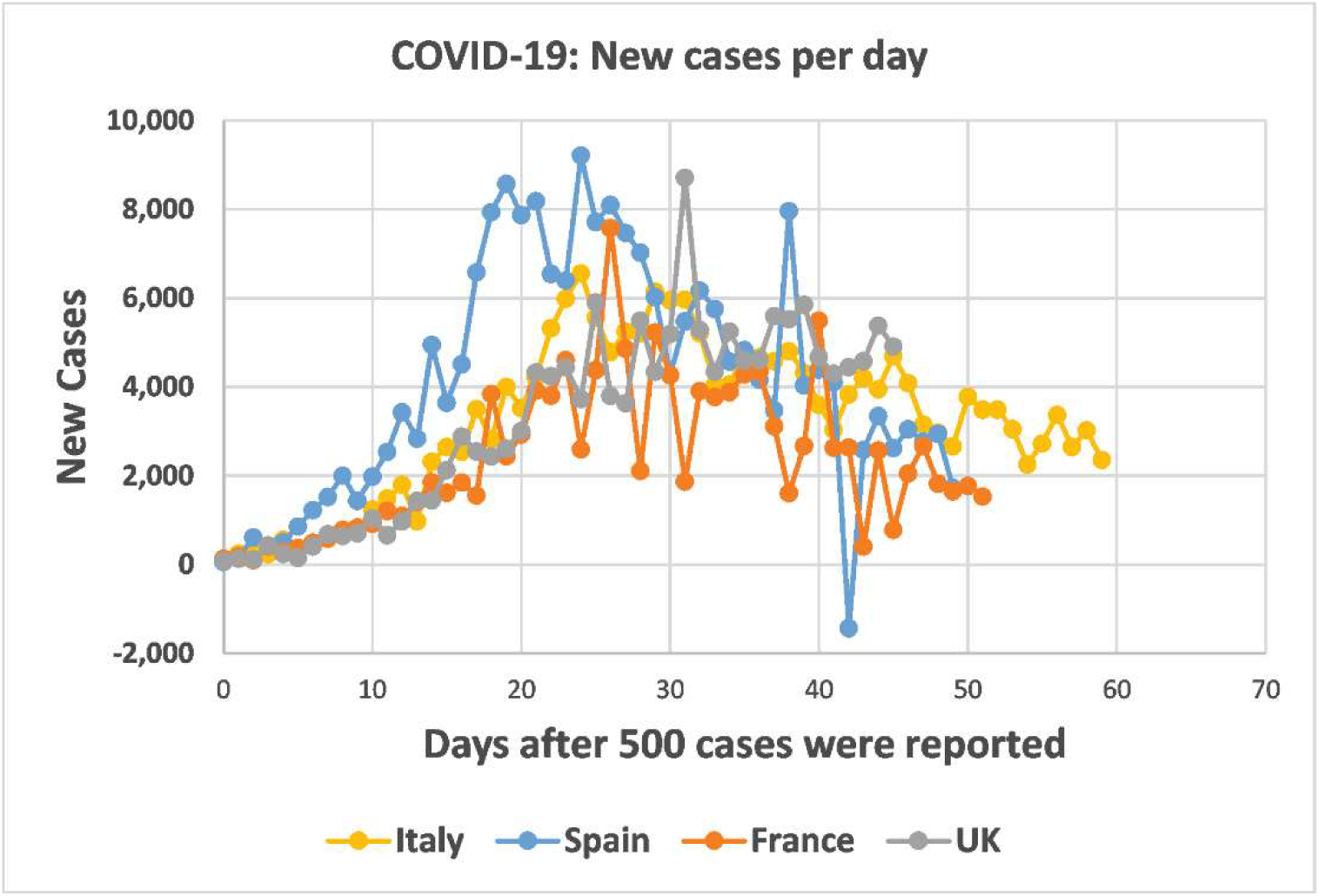
COVID-19 virus epidemics in Italy, Spain, France and UK: New reported cases up to April 26, 2020, as a function of days after the day that 500 cases were reported.

These graphs show that in all countries, except Greece, the growth of the epidemics is similar for the first approximately 10 days after the day that the number of infected persons reached 500. However, following this period, the behaviour of the epidemics is different, presumably reflecting the type of measures and the time that these measures were implemented, in each country. According to our definition, China reached the plateau around March 2, 2020 and South Korea around April 13, 2020.

The mathematical modelling of epidemics has a long and illustrious history; it began with the Kermack-McKendrick model, introduced in 1927 [5]. In this pioneering paper the population is divided into Susceptible, Infectious, and Recovered (removed) sub-populations. Then, specific ordinary differential equations are formulated specifying the time evolution of the functions representing these populations. The above work was certainly ahead of its time. It was rediscovered in the 1980s, and since then, it has provided the basis for a variety of deterministic models, known as SIR models. In particular, rigorous mathematical results for such models are derived in [6]. The extension of SIR to models involving partial differential equations is presented in [7]. Statistical models have also been highly effective for modelling aspects of epidemics.

In what follows we first postulate a general model for the accumulative number *N(t)* of individuals reported at time *t* to be infected by a viral epidemic. We *assume* that the function *N(t)* satisfies the ordinary differential equation

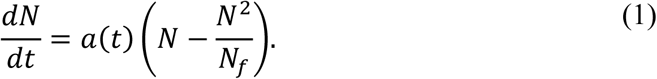

This is a Riccati equation that is specified by the time dependent function *α(t)* and the constant parameter *N_f_*. The function *α(t)* and the parameter *N_f_* depend on the basic characteristics of the particular virus and on the cumulative effect of the variety of different measures taken by the given country for the prevention of the spread of the viral infection. The dependence of *α(t)* on time reflects various time-dependent factors, including the fact that the effect of the different measures taken by the government depends on *t*. The case of *α(t)=constant* can be considered as an ‘ideal’ case.

Remarkably, although (1) is a nonlinear equation depending on time-dependent coefficients, it can be solved in closed form. Its solution depends on *α(t)*, the constant parameter *N_f_*, and the constant of integration *β*:

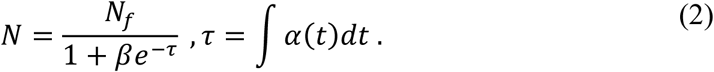

In the particular case that *α(t)* is a constant denoted by κ, equations (2) yield the classical logistic formula

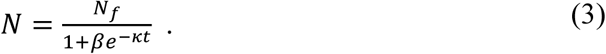

Interestingly, this simple formula is adequate for capturing the evolution *N(t)* of typical viral epidemics. For example, determining the three constant parameters *κ*, *β*, and *N_f_* of equation (3) with data from the Ebola virus epidemic of 2014 in Guinea, we find the excellent fit depicted in figure 3. Importantly, the above parameters remain essentially unchanged if we use a *smaller set* of data for their determination, which shows that the logistic model could also have been used for predictive purposes. *Throughout this paper the unknown parameters are determined by employing an error-minimizing algorithm described in section 2.1*. The cases of Liberia and Sierra Leone are very similar with the case of Guinea (the relevant data were obtained from the official site of the Centers for Disease Control and Prevention (CDC); they are official World Health Organization (WHO) data [8]).

**Figure 3.**
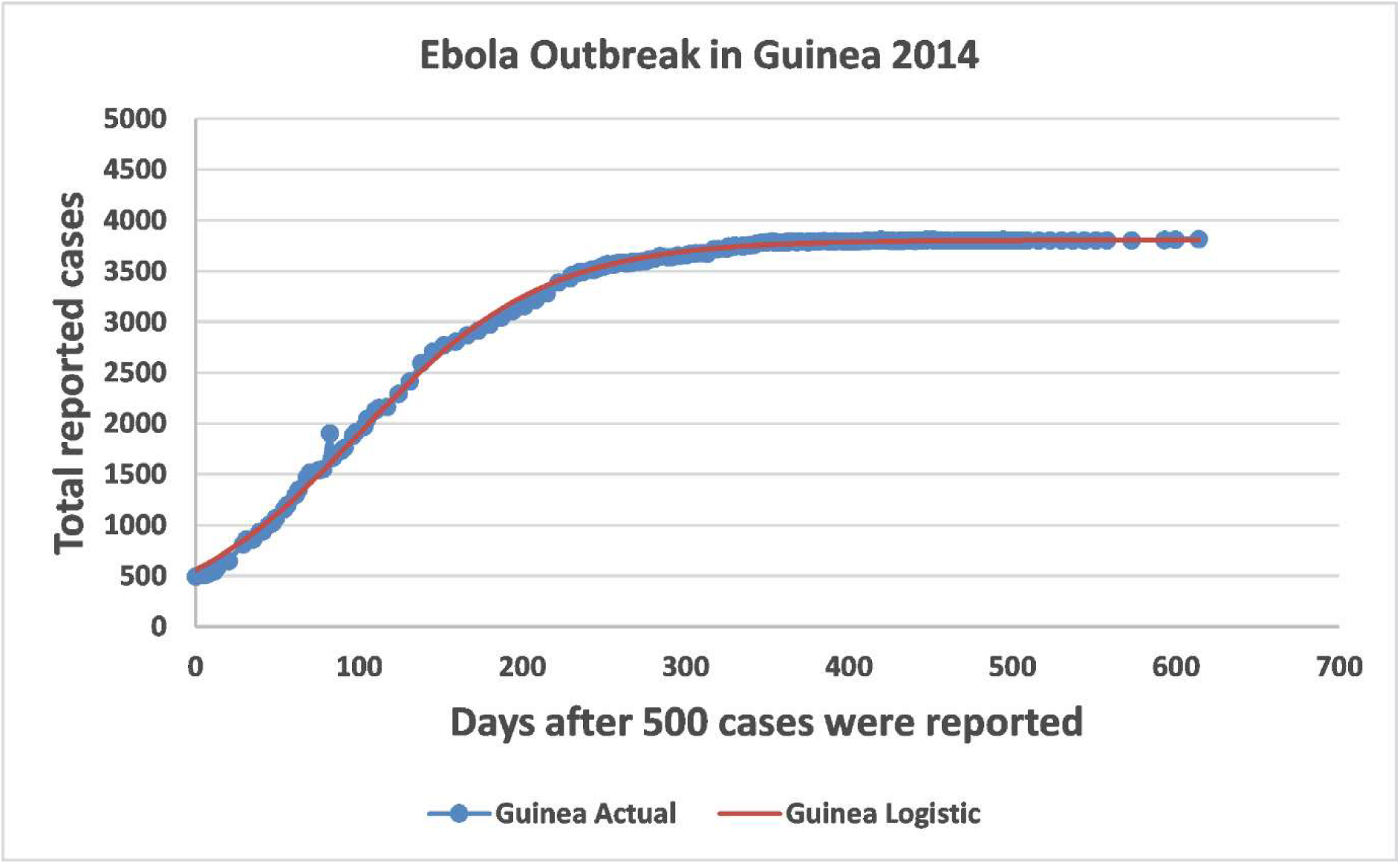
Ebola outbreak of Guinea in 2014: Predicted vs Actual for the total cumulative number of individuals reported to be infected, as a function of days after the day that 500 cases were reported. The logistic formula given by equation (3) gives an excellent fit for the actual data.

As it will be shown in section 3, the simple formula (3) also provides a good fit of the COVID-19 pandemic. However, the long series of existing data of the epidemics of China and South Korea shows that the logistic model does *not* provide accurate predictions. For example, figure 6a shows that if we use a subset of the existing data of the epidemic in South Korea for the determination of the parameters of the logistic model, and then compare the resulting graph of *N(t)* with the available data, we find that the logistic model *underestimates N(t)*. Thus, the logistic model provides a *lower bound* of the actual *N(t)*. This raises the following natural questions: first, is it possible to find a model yielding more accurate predictions than the logistic model and second, is it possible to construct a model that *overestimates N(t)*, which would then provide an *upper bound* of the actual *N(t)?* After experimenting with more than 50 different forms of *α(t)*, we have obtained affirmative answers to both of the above questions. We have introduced two novel models which will be referred to as *rational* and b*irational*. In the former model, the exponential function appearing in equation (3) is replaced by a *rational* function; in the birational model, the values of the parameters specifying this rational function change, depending on whether *t* is larger or smaller than a parameter denoted by *X*.

In order to evaluate the above two novel models and to establish their capacity for quantitative predictions in comparison to the logistic model, we implemented the following steps: (i) For the epidemics of China, South Korea, Italy, Spain, and France we computed the value of T. This value shows that for the above countries, the date of April 26, which is the last day that we have analyzed the data for *N(t)* and for its derivative, corresponds, respectively, to T+77, T+51, T+26, T+24, and T+22. (ii) For the epidemics of China and South Korea, we fitted the data for *N(t)* with the logistic, rational, and birational models. In addition, for China we plotted the relevant fit for the derivative of *N(t)*. From theses graphs it became clear that after *t* around 59 days (March 20, 2020), the effect of a second wave of reported infections became apparent. Thus, for the epidemic of China we limited our analysis until t=59 (thus, we fitted the three models using data only until t=59). (iii) In order to establish the predictive capacity of the rational and birational models in comparison to the logistic model, we determined the parameters of these three models for the epidemic in South Korea by using data *only* up to *t*=37. This date corresponds to the date of April 26 for Spain, and it differs only by two days with the date that corresponds to France and Italy. The rational and birational model provided, respectively, a *lower and upper bound* of the actual *N(t)*. Furthermore, the rational model gave a *better* lower bound than the logistic model. The analogous results for China are discussed in section 3. This analysis provides support of our speculation that the birational model can be used for *predicting* an upper bound for the actual *N(t)*. Using data until April 26, 2020, we found the following dates and number of reported cases may provide upper bounds for the characteristics of the plateaus in Italy, Spain, and France: June,14, 2020 (244,199 reported individuals), May 19, 2020 (225,267 reported individuals), May 20, 2020 (138,151 reported individuals).

## 2. The Basic Model

Let *F* denote the *relative infectivity* of the epidemic, defined by

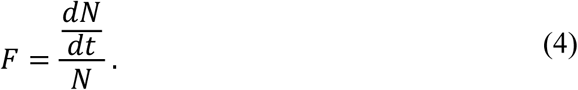

We *assume* that *F* is a *linear, time-dependent function of N*. Let the constant *N_f_* denote the final cumulative number of individuals reported to be infected. Taking into consideration that *F* vanishes when *N =N_f_*, we have

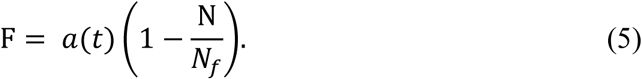

Inserting equation (5) in the definition (4) we find the basic equation modelling this situation, namely the Riccati equation (1).

The particular case of a Riccati equation with constant coefficients (corresponding to the case that *α* is constant) has appeared in a variety of dynamic processes, including the modelling of epidemics. Indeed, in the classical SIR model, mentioned in the introduction, if one assumes that *R=0*, then after replacing in the first order differential equation satisfied by *I, S* with *I-T*, where the constant *T* denotes the total population, one finds a Riccati equation of the form (3), where *α(t)* is replaced by a *constant*. Another notable example of the appearance of a constant coefficients Riccati equation in the mathematical modelling of infectious processes, can be found in the paper of Anderson and May [9]; this work describes the dynamic interaction of parasites with their host-environment. In this paper, whose impact in the field of mathematical biology was far reaching [10], a Riccati equation is formulated that involves a *single constant* parameter. It should be noted that certain optimal features of the dynamics of the SIR model are also characterised by a Riccati equation that involves three independent functions of time [6].

In order to solve the Riccati equation (1) we first use the independent change of variables specified by the second of equations (2). This gives rise to an equation similar to equation (1), where *α* is replaced by *1* and *t* by *τ*. This constant coefficients Riccati equation can be linearized via the change to the dependent variable

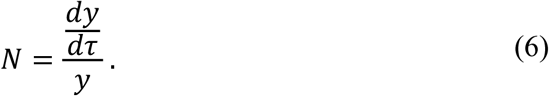

Indeed, substituting equation (6) into the above constant coefficients Riccati equation and simplifying, we find

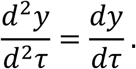

Solving this equation, substituting the resulting expression in equation (6), and simplifying, we find the first expression in equation (2).

We expect that the validity of the above model improves as *t* increases. Thus, we avoid evaluating equation (6) at *τ=0* to express *β* in terms of *N_f_* and *N* at *τ=0*. Instead we determine *β* by matching the expression obtained from the first of equations (2) with the actual data.

An important information provided by the above model is the time *T* when the maximum rate of infection is achieved: computing the second derivative of the right-hand side of the first of equations (2), and requiring that the resulting expression vanishes, we find that *T* satisfies the equation

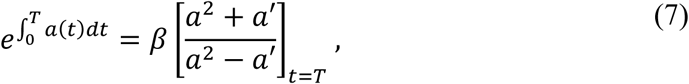

where throughout this paper prime denoted differentiation with respect to time. Using the above expression in the exponential occurring in the expressions for *N* and *N′* evaluated at *t=T*, we find

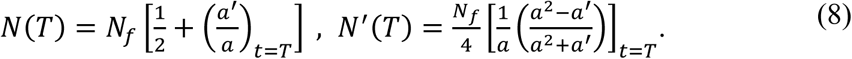

For the logistic model we have *α(t) = κ*. Thus, equations (8) yield

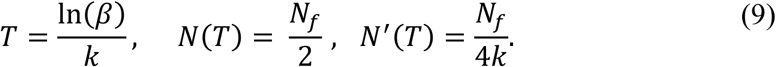

Taking into consideration that the logistic model is a good approximation of the relevant dynamic process, the above value of *T* provides an *approximate* value for the time that the inflection point is reached.

The rational and birational models are defined, respectively, as follows:

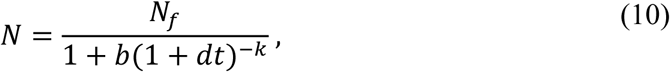

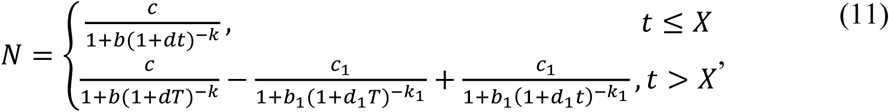

with *X* in the neighbourhood of *T*. The birational model is based on the natural assumption that the parameters of the rational function specifying the function *N(t)* are different before and after *T*. It is quite satisfying that this very simple model yields the best fits among more than 50 models that were tested. Among those models were several ‘fractal’ models (in the simplest such model, the exponent k·t in the logistic formula was replaced with k·t^μ^).

Letting in equation (11) *t* → ∞ we find:

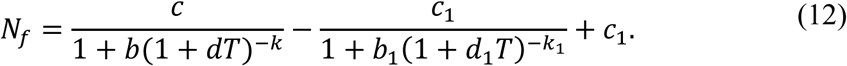

By comparing equations (10) and (11) with the first of equations (2), it is straightforward to determine *α(t)* for both the rational and the birational models: for the rational model

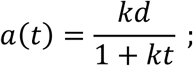

for the birational model

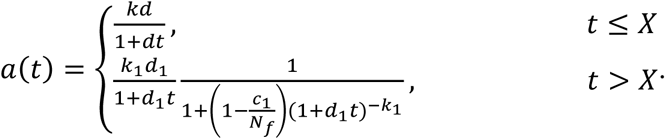

If *b, c, d, k*, are close to *b_1_, c_1_, d_1_, k_1_*, then *N_f_* is close to c_1_, and hence the value of *α(t)* after *X* is close to the value of *α(t)* before *X*.

Computing the second derivative of the right-hand side of equation (10) and equating the resulting expression to zero, we find that the value of *T* for the rational model is characterized by the equation

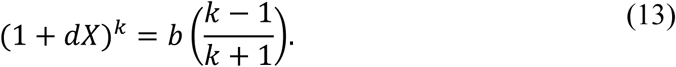

Similarly, for the birational model where the parameters *b, d, k*, are replaced with *b_1_, d_1_, k_1_*, respectively.

Replacing the rational function in the expressions for *N* and of its derivative with the rational function of equation (13), we find that for the rational model

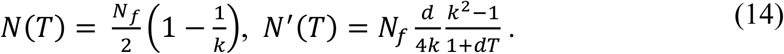

Similar expressions are valid for the birational model.

### 2.1 Optimization Method

We obtained the time-series data for the Coronavirus Disease (COVID-19) for China, South Korea, France, Spain, and Italy from the official site of the European Centre for Disease Prevention and Control [11]. We arranged the data in the form of individuals *N* reported to be infected over time measured in days, after the day that the number of cases reached 500.

All evaluated models were fitted using the *simplex algorithm*, which is an iterative procedure that does not need information regarding the derivative of the function under consideration. The algorithm creates a ‘random’ simplex of *n + 1* points, where *n* is the number of the model parameters that need to be estimated. The simplex changes iteratively by reflection, expansion, and contraction steps, until it finds the model parameters that minimize the given likelihood function. The constrained variation of the simplex algorithm [12, 13] available in MATLAB® was used for all tested models; an *L1-norm* was employed in the likelihood function to improve robustness [14]. The simplex algorithm is particularly effective for cases where the gradient of the likelihood functions is not easy to calculate. Random parameter initializations were used to avoid local minima. The simplex algorithm was chosen because it performed better than the nonlinear least-squares curve fitting algorithms evaluated in this work, namely the Levenberg-Marquardt [15] and the trust-region-reflective [16] algorithms.

The stability of the fitting procedure was established by using the following simple criterion: different fitting attempts based on the use of a fixed number of data points, must yield curves which have the same form beyond the above fixed points.

The fitting accuracy of each model was evaluated by fitting the associated formula on *all* the available data in a specified set. The relevant parameters specifying the logistic, rational, and birational models are given on table 1.

**Table 1.** Model parameters for the different models for China, South Korea, Italy, Spain and France.

|  |  | China | South Korea | Italy | Spain | France |
| --- | --- | --- | --- | --- | --- | --- |
| Logistic model | $N_f$ | 80,958 | 10,681 | 191,473 | 201,799 | 124,286 |
| | $k$ | 0.2266 | 0.1087 | 0.1257 | 0.1681 | 0.1489 |
| | $b$ | 75.1210 | 4.0870 | 61.1088 | 71.0470 | 77.0586 |
| | $T$ | 19 | 13 | 33 | 25 | 29 |
| | $R^2$ | 0.9968 | 0.9516 | 0.9955 | 0.9971 | 0.9984 |
| | $RMSE$ | 1814 | 687 | 4649 | 4186 | 1826 |
| Rational model | $N_f$ | 81,755 | 11,275 | 237,876 | 232,426 | 142,411 |
| | $k$ | 11.9175 | 2.0166 | 3.2074 | 3.9584 | 4.9767 |
|  | ***b*** | 136.8769 | 36.1329 | 6,852.7948 | 1,485.8026 | 528.9068 |
|  | ***d*** | 0.02700 | 0.3982 | 0.3993 | 0.1962 | 0.08124 |
|  | ***R*<sup>2</sup>** | 0.9962 | 0.9924 | 0.9995 | 0.9996 | 0.9996 |
|  | ***RMSE*** | 2015 | 278 | 1655 | 1564 | 979 |
| **Birational model** | ***k*** | 4.3787 | 4.8476 | 6.2579 | 8.7412 | 7.0324 |
|  | ***b*** | 497.7721 | 35.6142 | 426.8576 | 268.5638 | 488.6477 |
|  | ***c*** | 79,423 | 9,290 | 187,326 | 170,536 | 145,396 |
|  | ***d*** | 0.16114 | 0.1110 | 0.05228 | 0.03925 | 0.04672 |
|  | ***k*<sub>1</sub>** | 6.0242 | 2.4669 | 4.2406 | 5.2741 | 6.9915 |
|  | ***b*<sub>1</sub>** | 462.1967 | 39.9082 | 400.1164 | 140.2595 | 614.7524 |
|  | ***c*<sub>1</sub>** | 119,053 | 7,332 | 210,077 | 229,593 | 108,744 |
|  | ***d*<sub>1</sub>** | 0.10685 | 0.1632 | 0.07052 | 0.05667 | 0.04306 |
|  | ***R*<sup>2</sup>** | 0.9971 | 0.9973 | 0.9999 | 0.9997 | 0.9994 |
|  | ***RMSE*** | 1762 | 164 | 697 | 1296 | 1132 |

**Table 2.** Plateau characteristics as determined by the different models.

|  | **China** |  | **South Korea** |  | **Italy** |  | **Spain** |  | **France** |  |
| --- | --- | --- | --- | --- | --- | --- | --- | --- | --- | --- |
|  | Plateau (days) | Plateau Cases | Plateau (days) | Plateau Cases | Plateau (days) | Plateau Cases | Plateau (days) | Plateau Cases | Plateau (days) | Plateau Cases |
| **Actual** | 41 | 80,134 | 51 | 10,537 | - | - | - | - | - | - |
| **Logistic** | 38 | 79,866 | 47 | 10,424 | 68 | 189,232 | 53 | 199,877 | 59 | 122,834 |
| **Rational** | 41 | 80,229 | 50 | 10,454 | 102 | 227,961 | 72 | 225,267 | 75 | 138,151 |
| **Birational** | 41 | 79,885 | 56 | 10,653 | 108 | 244,199 | 71 | 226,424 | 72 | 143,428 |

## 3. Results

We first computed the inflection point for China, South Korea, Italy, Spain, and France. This occurred respectively, at t=19, t=13, t=33, t=25, and t=29 (see table 1). This corresponds to February 2, March 6, March 31, April 2, and April 4, 2020, respectively. For computing T, we require the time that the derivative of *N* becomes maximum. For this purpose, we used the model with the best fit in the neighborhood of the inflection point, which in all cases turned out to be the birational model.

Figures 4 and 5 show that the logistic, rational, and birational formulas provide accurate fits for the available data for the epidemics of China and South Korea. However, by carefully scrutinizing the situation of the China epidemic it became clear that after around t=59 the effect of a second wave of reported infections begins to have an effect. Thus, whereas for South Korea we used all the data up to our cut of date of April 26, 2020, to fit the 3 models, for China we used data only until t=59.

**Figure 4.**
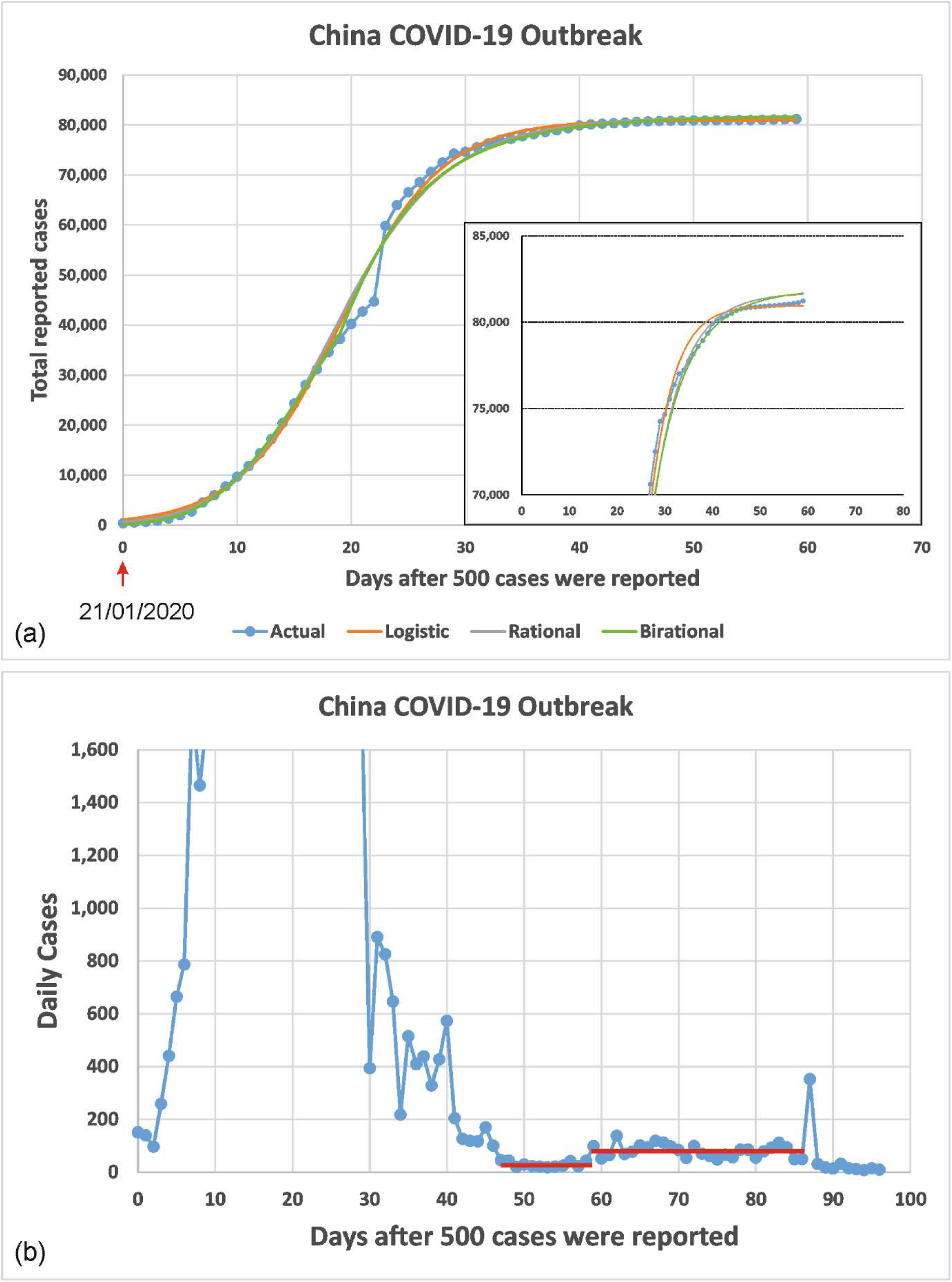
COVID-19 virus infection for China: (a) Predicted vs Actual for the total cumulative number of individuals reported to be infected as a function of days after the day that 500 cases were reported. The three models were fitted using data only up to t=59. The inflection point occurred at t=19 which corresponds to March 2, 2020. (b) Daily new cases as a function of days after the day that 500 cases were reported. This curve clearly indicates the impact of a second wave of reported infections after around t=59.

**Figure 5.**
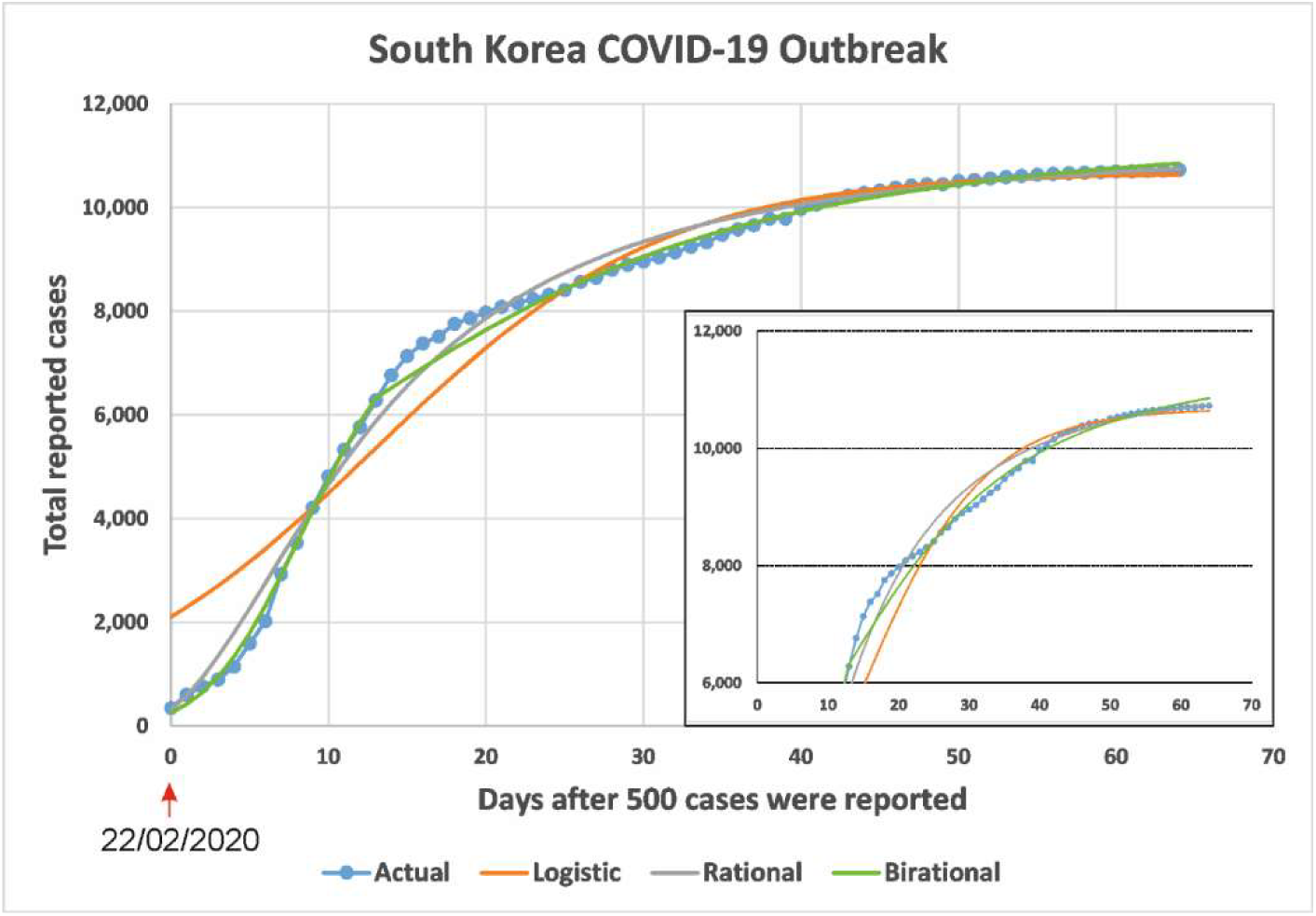
COVID-19 virus infection for South Korea: Predicted vs Actual for the total cumulative number of individuals reported to be infected as a function of days after the day that 500 cases were reported. The three models were fitted with data up to t=64 which corresponds to April 26,2020. The inflection point occurs at t=13 which corresponds to April 13, 2020.

It is important to emphasize that whereas each of the three equations (3), (10), and (11) can fit the data quite well, the predictive capacity of these formulas is *not* the same. This is best illustrated by using the epidemic of South Korea: figure 6a shows the fits of the above equations using data *only* up to t=37, which corresponds, approximately, to the date of April 26, 2020, of the epidemics of Italy, Spain and France. It is clear that each of the above equations gives a different curve. Furthermore, the rational and birational models provide a lower and upper bound of the actual *N(t)*, and the rational model is a better lower bound that the logistic formula. Regarding the birational model, we have experimented with different values of *X*. For all such values, the birational curve was above the rational curve; however, in order to obtain a curve that is above the actual *N(t)*, it was necessary to choose *X=T+9*. Since it is important to have an upper bound, we propose the following strategy for the choice of X: choose the value of *X* in the vicinity of T which yields the uppermost curve.

**Figure 6.**
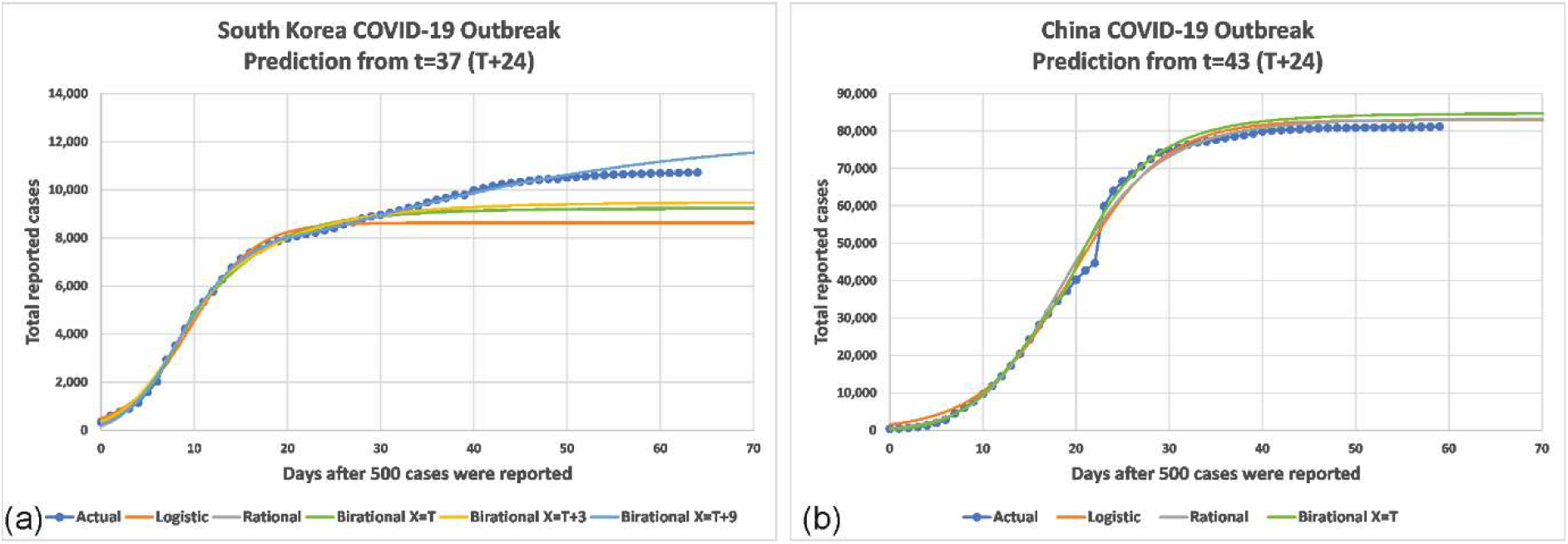
Predicted vs Actual for the total cumulative number of individuals reported to be infected as a function of days after 500 cases were reported for South Korea and China. The fits were obtained using data only up to t=37 and 43 respectively. These times correspond approximately to the date of April 26, 2020 for Italy, Spain and France.

The analogous results for China are shown in figure 6b. Now, since t=43 is already in the stable sigmoidal part of the curve of the actual data, the three curves corresponding to the logistic, rational and birational models are very close to each other and to the curve of the actual data. Importantly, the birational curve is above the curve of the actual data; thus, the birational model, as with the case of the South Korea provides an upper bound for *N(t)*.

Figure 7 presents the actual vs. predicted cumulative number of individuals reported to be infected with the COVID-19 virus as a function of days after 500 cases were reported, for Italy, Spain, and France. The three models were fitted with data up to April 26, 2020, which corresponds respectively to T+26, T+24, and T+22. For the epidemic of Italy, the logistic model predicts a plateau on May 5, 2020 (day 59 after the day that 500 cases were reported) with 189,232 reported individuals; the rational model predicts a plateau on June 8, 2020 (day 102) with 227,961 reported individuals; and the birational model predicts a plateau on June 14, 2020 (day 108) with 244,199 reported individuals. Clearly, again, the logistic model underestimates the actual plateau day and the number of cases infected, since on April 26, 2020 (last day of acquired data for this study) the number of infected cases for Italy had reached 195,351. For the epidemic of Spain, the logistic model predicts a plateau on April 30, 2020 (day 53 after the day that 500 cases were reported) with 199,877 reported individuals; the rational model predicts a plateau on May 19, 2020 (day 72) with 225,267 reported individuals; and the birational model predicts a plateau on May 18, 2020 (day 71) with 226,424 reported individuals. Again, the logistic model underestimates the actual plateau day and number of cases infected, since on April 26, 2020 the number of infected cases for Spain had reached 207,634. For the epidemic of France, the logistic model predicts a plateau on May 4, 2020 (day 59 the day that 500 cases were reported) with 122,834 reported individuals; the rational model predicts a plateau on May 20, 2020 (day 75) with 138,151 reported individuals; and the birational model predicts a plateau on May 17, 2020 (day 72) with 143,428 reported individuals. Again, the logistic model underestimates the actual plateau day and number of cases infected, since on April 26, 2020, the number of infected cases for France had reached 124,114.

**Figure 7.**
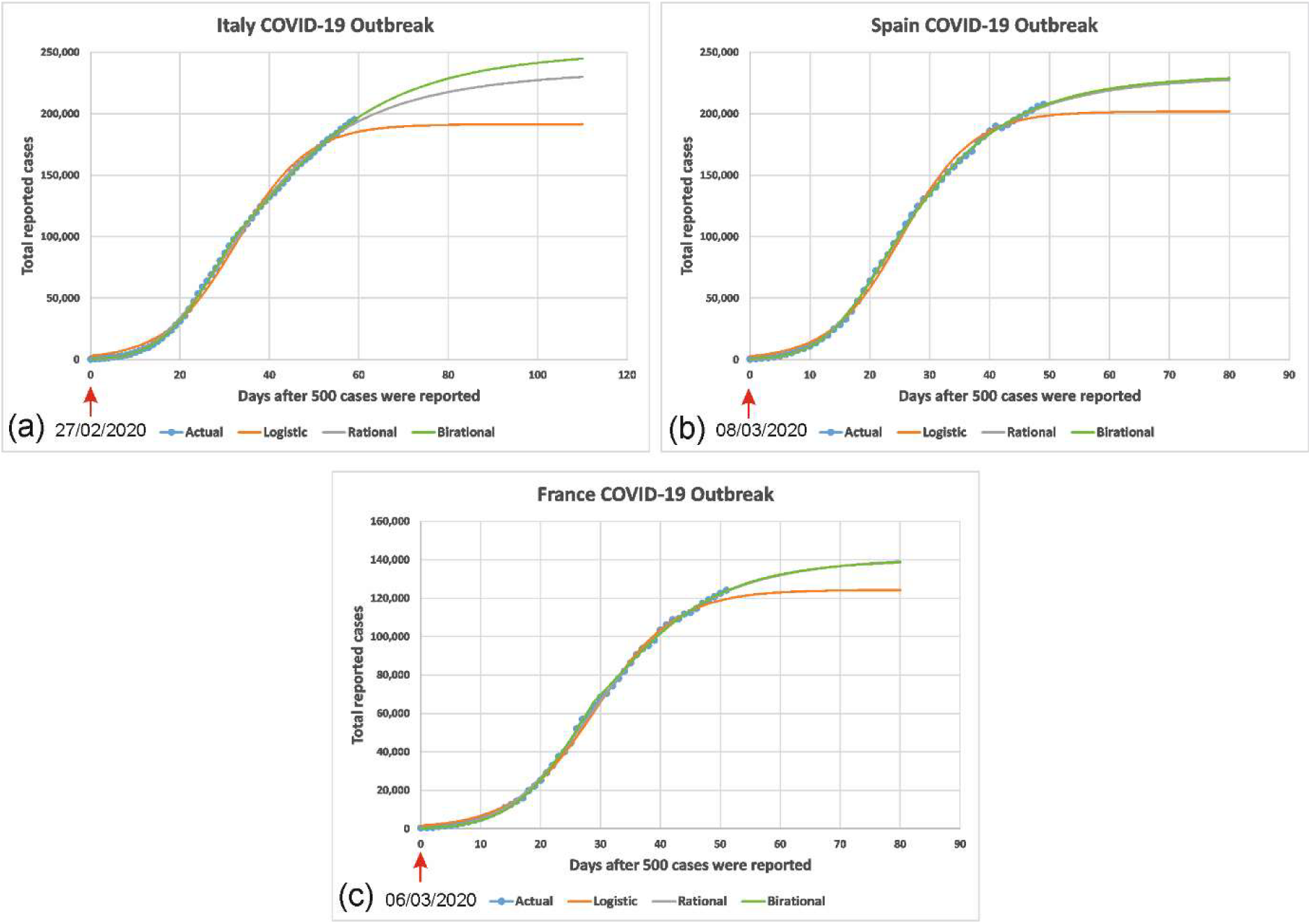
COVID-19 virus infection for Italy, Spain and France: Predicted vs Actual for the total cumulative number of individuals reported to be infected as a function of days after 500 cases were reported. The three models were fitted with data up to April 26, 2020. This corresponds to t=59 data points for Italy, t=49 data points for Spain and t=51 data points for France. The inflection point occurs on March 31, 2020, for Italy, on April 2, 2020, for Spain and on April 4, 2020, for France.

Regarding the birational model, for the cases of Italy and Spain the uppermost curves were obtained by using *X=T*, whereas for France for *X=T-3*.

## 4. Conclusions

Several useful models elucidating aspect of the COVID-19 pandemic have already appeared in the literature; they include the following: (i) A model for simulating the transmissibility of COVID-19 from bats to humans is presented in [17]. (ii) The calculations of exponential growth and maximum likelihood are used in [18] to determine the reproductive number of 2019-nCoV and SARS in China. (iii) The formulation of a Susceptible-Infected-Recovered-Dead (SIDR) model, together with the knowledge of data from China in the period January 11 to February 10, 2020, is used in [19] to estimate the associated per day infection mortality and recovery rates. (iv) In [20], by combining a stochastic model for the COVID-19 infection with the knowledge of data from China during January and February 2020, the probability that newly introduced cases might generate new outbreaks is calculated. (v) In [21], an SIDR model supplemented with mean-field kinetics is used to calculate the time and peak of confirmed infected individuals in China, Italy and France. (vi) In [22], the effect of social distancing was studied by using a model where the population was divided into those who are asymptomatic or have mild symptoms (95.6%), those who are hospitalized but do not require critical care (3.08%), and individuals who require critical care (1.32%); seasonal variations were incorporated by allowing the basic reproduction number to be a time-dependent function following a cosine curve that peaks in early December.

Here, we have modelled the cumulative number *N* of persons reported to be infected by COVID-19 in a given country as a function of time, in terms of the Riccati equation (1). Although this equation is a nonlinear ordinary differential equation containing time dependent coefficients, it was solved in closed form, yielding (2). For appropriately chosen functions *α(t)*, the first of equations (2) provides a flexible generalization of the classical logistic formula that has been employed in a great variety of applications, including the modelling of infectious processes.

The fact that *α* is now a function of *t* has important implications. In particular, it made it possible to construct the rational and birational models, which at least for the epidemics of China and South Korea provide, respectively, were able to provide more *accurate predictions*. Furthermore, the birational model provided an *upper bound of the actual N(t)*. This suggests that the methodology introduced in our work may be used for providing bounds of important aspects of the current pandemic. For example, it can provide reasonable estimates for the time that the plateau will be reached as well as the number of persons that will be reported to be infected at that time.

The approach presented here has several novel and useful features: (i) For the case that an infection that has been stabilized, any of the three models analyzed here can be used for the evolution of the cumulative number of persons reported to be infected, via a simple analytical expression. This expression can be used for a variety of purposes. (ii) More importantly, the rational and birational models can be used for *predictive* purposes, providing accurate estimates for the characteristics of the plateau. (iii) Our approach has the capacity for increasing continuously the accuracy of the predictions: as soon as the epidemic in a given country passes the time *T*, the rational model can be used; furthermore, when the sigmoidal part of the curve is approached, the rational model can be supplemented with the birational model (a simple criterion of checking whether the birational model can be used is given in section 2.1). Also, as more data become available, the parameters of the rational and of the birational models can be re-evaluated; this will yield better predictions. (iv) The Riccati equation (1) together with the flexibility of the arbitrariness of *α(t)*, offer the possibility of deciphering basic physiological mechanisms dictating the evolution of *N(t)*. In particular, following the transient stage of the epidemic, it may be envisioned that *α(t)* becomes a function of *N* instead of a function of t. By plotting *α* in terms of *N* is possible to scrutinize a posteriori their relationship: we find that after *t* approximately equal to T-7 the relation between the relation between *α* and *N/500* is, remarkably, *linear*, see figure 8.

**Figure 8.**
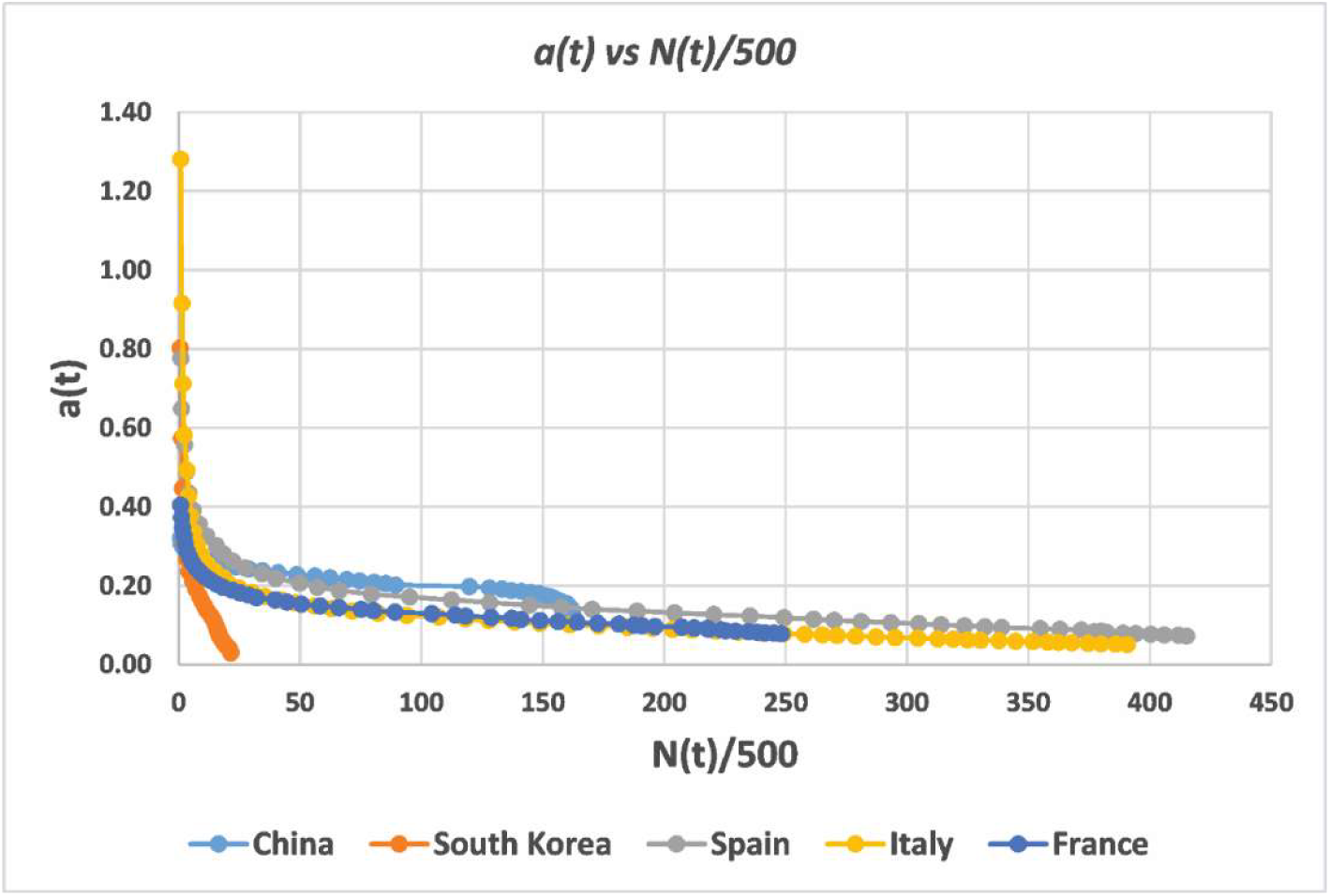
Plot of a(t) as a function of N(t)/500 for the COVID-19 virus infection for China, South Korea, Spain, Italy and France. After t around T-7, *a(t)* for China, Italy, Spain and France is the same linear function of N(t).

How can the apparent success of the simple approached followed in our work can be explained? Presumably, the constant *N_f_* defining equation (1), the constant of integration *β* entering the associated solution, and the constant parameters specifying the function *α(t)*, must capture the essence of the underlying time-evolution process. This suggests that the cumulative effects of a variety of different mechanisms express themselves via the few parameters entering in the explicit solution formulas (3), (10) and (11). In this connection, it is worth recalling that the *single* parameter characterizing the Riccati equation of the celebrated Anderson-May model mentioned earlier, represents the *cumulative effect* of different biological mechanisms: it is shown in [9] that this constant can be expressed in terms of the population size parameter H, the mortality rate of uninfected hosts parameter b, the mortality rate of infected hosts parameter *α + b*, and the rate at which infected hosts recover and become susceptible parameter *γ*.

An additional partial explanation of the success of our approach is that it shares the same philosophy employed by the powerful technique of Artificial Intelligence known as machine learning. Indeed, the explicit formulas (3), (10) and (11) used in this work can be thought of as ‘algorithms’, where given *t*, they predict *N;* these algorithms are characterized by several parameters, which are fixed by the ‘knowledge’ of the data. Thus, the more data are available, the better this algorithm ‘learns’ how to make accurate predictions. Hence, choosing these parameters by requiring that the analytical solution matches the data curve, is consistent with the approach of machine learning. This point is further explored in [N. Dikaios, A. S. Fokas, and G. A. Kastis, Deep Learning for Predicting Aspects of the COVAD-19 Pandemic, in preparation].

Taking into consideration the ubiquitous use of the logistic model and the fact that equations (10) and (11) provide variations of the logistic formula, these equations may be useful for the modelling of a variety of phenomena.

The fact that two different viral infections, namely COVID-19 and Ebola, are modelled by the same ordinary differential equation, suggests that the Riccati equation (1) proposed here plays a generic role in the modelling of viral epidemics.

It is worth noting that the so-called Burgers equation, which is an evolution partial differential combining the generic effects of diffusion and nonlinear convection, admits a travelling wave solution that satisfies the Riccati equation (1) in the case that *α(t)* is a constant (this constant specifies the speed of propagation of the traveling wave, whereas *N_f_* is free parameter appearing in Burgers’ equation). Hence, the mathematical analysis presented in this work may also be relevant for some of the phenomena modelled by appropriate generalizations of the Burgers equation.

## Data Availability

We obtained the time-series data for the Coronavirus Disease (COVID-19) for China, South Korea, France, Spain, and Italy from the official site of the European Centre for Disease Prevention and Control.

https://www.ecdc.europa.eu/en/geographical-distribution-2019-ncov-cases

## Data accessibility

Analytic spreadsheets of our fitting results will be available as part of the electronic supplementary material.

## Authors’ contributions

A.S.F. conceived, designed and supervised the study. N.D. and G.A.K. performed the fitting experiments and analysis of the results. A.S.F. wrote the initial draft of the manuscript. All authors critically revised and improved the manuscript in various ways. All authors reviewed the manuscript and gave final approval for publication.

## Funding

A.S.F. acknowledges support from EPSRC via a senior fellowship.

## Acknowledgments

A.S.F is grateful to V. Marmarelis, whose deep insight regarding the remarkable range of the applicability of the Riccati equation provided the impetus for this work. He also thanks K. Kalimeris and Y. C. Yortsos for useful suggestions.

## Notes

### Competing Interest Statement

The authors have declared no competing interest.

### Funding Statement

No external funding was received.

